# Stochastic model for COVID-19 in slums: interaction between biology and public policies

**DOI:** 10.1101/2021.01.06.21249318

**Authors:** H.G. Solari, M.A. Natiello

**Affiliations:** Departamento de Física, FCEN-UBA and IFIBA-CONICET; Centre for Mathematical Sciences, Lund University

**Keywords:** stochastic compartmental model, viremic levels, surveillance protocols, Markov Jump process

## Abstract

We present a mathematical model for the simulation of the development of an outbreak of COVID-19 in a slum area under different interventions. Instead of representing interventions as modulations of the parameters of a free running epidemic we introduce a model structure that accounts for the actions but does not assume the results. The disease is modelled in terms of the progression of viremia reported in scientific works. The emergence of symptoms in the model reflects the statistics of a nation-wide highly detailed database consisting of more than 62000 cases (about a half of the confirmed by RT-PCR tests) with recorded symptoms in Argentina. The stochastic model displays several of the characteristics of COVID-19 such as a high variability in the evolution of the outbreaks, including long periods in which they run undetected, spontaneous extinction followed by a late outbreak and unimodal as well as bimodal progressions of daily counts of cases (second waves without ad-hoc hypothesis). We show how the relation between undetected cases (including the “asymptomatic” cases) and detected cases changes as a function of the public policies, the efficiency of the implementation and the timing with respect to the development of the outbreak. We show also that the relation between detected cases and total cases strongly depends on the implemented policies and that detected cases cannot be regarded as a measure of the outbreak, being the dependency between total cases and detected cases in general not monotonic as a function of the efficiency in the intervention method. According to the model, it is possible to control an outbreak with interventions based on the detection of symptoms only in the case when the presence of just one symptom prompts isolation and the detection efficiency reaches about 80% of the cases. Requesting two symptoms to trigger intervention can be enough to fail in the goals.

## 1 Introduction

Ever since the emergence of COVID-19 [1], mathematical models have been proposed to examine, illustrate and forecast the possible evolution of the pandemic, as well as recommending public measures for managing it. Modelling epidemics has to deal with a variety of difficulties at different levels and the present pandemic is not an exception.

This work adopts a stochastic approach –following a line of thought developed over the last century [2, 3, 4, 5]– that rests on counting populations in its natural form and evolving their numbers at characteristic events. In relation to them, ODE models represent the evolution of average fractions of populations in a large-population limit [6, 7, 8, 9] ^1^. In this respect, one of our aims is to explore, at least within the limited scope of the situation considered, the relevance of stochasticity in our perception of the pandemic.

A good part of the literature has addressed the phenomena of asymptomatic carriers of SARS-CoV-2 [11, 12, 13, 14, 15, 16, 17]. Unfortunately, the label “asymptomatic” has been used with different meanings, going from ‘not presenting the expected symptoms at the moment of infecting someone else’, as in [11, 13] to “never, in the course of the infection, presenting symptoms” [18]. In all cases, asymptomatic and pre-symptomatic are considered as objective categories pertaining to the relation between the infected person and the infectious agent. The influence of the actions of the public health system and the perception of illness by the patients in building these categories has not been properly examined, thus preventing any improvement of these actions.

In previous modelling work either asymptomatic carriers of SARS-CoV-2 have not been considered or they have been incorporated using an ad-hoc hypothesis, such as that the ratio between asymptomatic and symptomatic cases is constant (see, e.g., [19]). In contrast, our model incorporates a detection component based in what it is known of detection policies. Another sharp difference with earlier work is that we model a variable contagiousness and not only a variable contagious period. Furthermore, intrinsic stochasticity is included in contrast with the extrinsic stochasticity (added a-posteriori ^2^) included in [19] and few other works. A search in PubMed^3^ with keywords *covid-19, mathematical, model* offered 540 articles. A refinement with keywords *covid-19, model, asymptomatic, stochastic* ends in six scientific publications (as of 2020-09-09) plus one news article (not a research article) for a specialized magazine. Of the later six, the reference [19] is the most closely related to our work, hence our decision to indicate only the differences of the present work with a related one among the pre-existing papers.

In this work we will take a complex systems view. We begin by acknowledging that the COVID-19 epidemic is no longer a free-running epidemic but rather one in which there is a strong interaction between the public health system and the population dynamics of the outbreaks. Changes in the evolution of an outbreak trigger changes in the consideration of which characteristics of the COVID-19 cases should (or should not) trigger public action. This indicates that there is a clear interaction between these systems and they cannot be considered independent. To illustrate the point we will use the various criteria of “COVID-19 case” used in our home country (HGS), Argentina, following recommendations by the World Health Organization (WHO). We will produce compartments that relate to the evolution of the case in medical or biological terms as well as to the categories corresponding to the different protocols to be applied to the case.

The response to an epidemic requires not only the mobilisation of public resources but the participation of the public as well. To organize the actions required for each individual case COVID-hot-lines and web-servers have been organized world wide. Such help services indicate which measures to take by those that suspect they are developing COVID-19, and prompt official actions if needed. Hospitals and health centres, as well as help services, are coordinated in their actions by protocols. A main tool of these protocols is the *suspected-case* criterion. This criterion regulates state intervention and depends on clinical symptoms of the (potential) patient and other circumstances. The criterion constitutes a difficult balance between the administration of resources (for example use of reverse transcription polymerase chain reaction (RT-PCR) kits and laboratories), the developmental stage of the epidemic, the mortality risk of the case and more. As in any decision taken under real circumstances (limited resources), establishing the suspected-case criterion implies trade-offs. When diagnostic resources –such as RT-PCR tests– are limited, a conflict emerges: should we reserve them for individual diagnosis (for example, to confirm diagnostic by symptoms in cases of doubt or concern) or perhaps use them in epidemiological surveillance (triggering actions such as contact tracing or sample pooling monitoring) as well? In any intermediate cases: in which proportions?

Should the general criterion depend on being a *contact* of a COVID-19 case? Does it make sense to require weaker symptoms for the population which is aware of having epidemiological contact with COVID-19 cases rather than for the *communitarian* cases that cannot account for how they could have been infected? Actually, it could make sense if by such measures we were able to achieve a more efficient use of a scarce resource to be reserved for diagnosing related to treatment (a private/individual criteria contrasting to public/epidemic criteria). The question must be put: is it correct to focus our attention in travellers and their contacts at the beginning of the outbreak? Is efficiency really boosted by requiring two relevant symptoms of a list for potentially communitarian cases and only one to people with epidemiological contacts? In the context of the propagation of SARS-CoV-2, what are the consequences of such decisions? We will address these questions implementing a model apt for answering them.

To set the grounds for our model, we analyse data collected by the Public Health Ministry of Argentina, made available to us through the COVID-19 initiative under the Ministry of Science and Technology. The model incorporates medical findings regarding the transmission of SARS-CoV-2 as well as actions taken by the health authorities and to a certain extent the social behaviour of the population. We apply the model to small slums (variously called in South-America: *villas miseria, villas de emergencia, cantegril, favelas*, etc.) where the conditions of homogeneous contact, frequently used to simplify the modelling task, are closer to be fulfilled. We show how the model predicts epidemic circulation below the detection level for surprisingly long periods of time. Also, we illustrate that “average epidemics” are not good representatives to grasp the dynamics, and that the undetected (mild, unrecognized, presymptomatic, “asymptomatic”) cases are in good proportion the result of public policies coupled to the characteristics of the illness. The outcome of three forms of surveillance and public action are comparatively analysed.

In Section 2 we describe the model, from its basis –supported in both biology and social behaviour all the way to the algorithm implementing a Markov Jump process [3, 4]. Results are presented in Section 3 and discussed in the following Section 4. Section 5 finally sums up the Conclusions.

## 2 The Model

### 2.1 Biological and social input

#### 2.1.1 What is a COVID-19 case?

We review the evolution of the definition of “case” along with the development of the pandemic. In many countries this definition emerges from the national Health authorities, following recommendations from WHO. By 27 January 2020 there were comparatively few cases outside China. Apart from special considerations for sanitary operators, the definition of *suspected case* from the Italian health authorities^4^ considered two situations: (A) severe acute respiratory infection (fever, cough and request for hospitalisation) and presence in risk zones a few days before the onset (at that moment mainly Wuhan/Hubei), or (B) acute respiratory infection and either recent presence at Wuhan live animal market or recent close contact with a *confirmed* (positive PCR test) or *probable* case (a PCR-tested suspected case without a conclusive result). By 22 February ^5^ severity and hospitalisation were no longer required for (A) and dyspnea was recognized among possible symptoms. By 9 March ^6^, the considered situations were three: acute respiratory infection (with at least one among fever, cough and difficulty in breathing) without other aetiology and either (A) recent presence in areas of local transmission of the disease or (B) close contact with probable or confirmed cases. The third situation considered (C) cases presenting severe acute respiratory infection (fever and at least one symptom of respiratory disease) requiring hospitalisation and without another aetiology that fully explains the clinical presentation. This new item acknowledges the existence of the illness regardless of any presence in risk zones or close contact with probable or confirmed cases.

Also the concept of *close contact* evolved during the period. By 31 January “risk contacts”^7^ considered only recent (within 14 days) travel or cohabitation with a COVID-19 patient (apart from special considerations for sanitary operators). The concept evolved to that of close contact, becoming highly detailed in what regards social distance (2m, 15 min) and hygiene already by 27 February 2020 ^8^.

At the end of May, specific instructions for *contact tracing*^9^ (already operative, though) had been developed.

The criteria for identification of cases shifts focus along the pandemic. At the beginning, the focus is in the “virus import” from other regions where it is active, while the local diffusion becomes relevant only some weeks/months later. The trade-off in the identification generates “classes” of contagion depending on the criterion.

Along with the case criteria, surveillance and control criteria are developed. At the beginning of the pandemic, *passive surveillance* (i.e., to wait for the spontaneous appearance of patients, except perhaps for travellers) was the most common attitude.Soon after, many countries developed different degrees of contact-tracing (with varying success), even revealing preexistent flaws in the various national health and care systems.

In Appendix A we show the evolution of the criteria in Argentina and its relation with Italy’s case.

The decision of what to consider a *suspected case*, and when further actions are to be taken, is a critical one. However, it is not clear which is the overall criteria, meta-criteria, adopted by Italy or Argentina, presumably upon recommendations of WHO. It appears that the meta-criterion is to keep an even level of certainty of being a COVID-19 case for each individual case. It is then pertinent to explore whether this goal is achieved or not and if such goal is epidemiologically sound.

We discuss this issue with data from Argentina^10^. In Table 1 we report PCR results for health workers after 6 June, from the data set of October 5th, 2020 with 62920 cases with symptoms information (29958 positive and 32962 negative)^11^. Health workers can be assumed to be more accurately monitored than other patient groups. At 6 June, the criterion for suspicious case for health workers was changed to presenting one symptom belonging to the set: fever, cough, anosmia, dysgeusia, dyspnea, odynophagia (see Appendix A). In 4 August 2020, the set of symptoms was extended to headache, diarrhoea and vomits.

**Table 1:** Health workers. Number of confirmed PCR-positive and negative cases displaying at least one or two symptoms (1*s*, 2*s*) from the set given in the text. *F* +: Cases with fever plus another symptom. *H*: cases requiring hospitalisation. Positivity odds in data set are 0.9089. *P*_*X*_ (±) stands for the probability of having one symptom or more of SARS-CoV-2 being positive (negative) for each category. Odds ratios OR are the ratio of the odds under the symptoms condition to the odds in the full set.

| PCR | Health workers |  |  |  |  |  |  |  |
| --- | --- | --- | --- | --- | --- | --- | --- | --- |
| | $\geq 1\text{ s}$ | $P_{1s}(\pm)$ | $\geq 2\text{ s}$ | $P_{2s}(\pm)$ | $F+$ | $P_{F+}(\pm)$ | $H$ | $P_H(\pm)$ |
| Pos | 29465 | 0.9835 | 23688 | 0.7907 | 13415 | 0.4478 | 2814 | 0.0939 |
| Neg | 32290 | 0.9796 | 23347 | 0.7083 | 9015 | 0.2735 | 2299 | 0.0697 |
| OR | 1.0040 |  | 1.1163 |  | 1.6373 |  | 1.3467 |  |
| Tot | 61755 | 0.9815 | 47035 | 0.7475 | 22430 | 0.3565 | 5113 | 0.0813 |

For health workers, 98% of the cases that reported symptoms ^12^ presented at least one symptom in the extended set. Among them, 48% were diagnosed as COVID-19 cases using RT-PCR. Considering cases reporting at least two symptoms, the number of cases falls by 23% but the positive cases within the group move up only to 50%. If the criterion is “fever and one symptom”, the case fall is 64% while the positivity within this smaller set raises to 60%. Similar trends are found for the whole patient data set.

The data indicates that requiring more symptoms results in missing positive cases. The improvement in positivity rates is outnumbered by the large or very large fall in detected cases, with no significant improvement in the use of resources. At the early stages of the epidemic only hospitalized patients with pneumonia were considered as possible COVID-19 cases. In such case the detection ability drops to less than 10% of the cases showing symptoms.

#### 2.1.2 Viremia, symptoms and contagiousness

An important ingredient of any model concerning the evolution of the disease requires the description of a contagion mechanism at the individual level. It is important to relate when, how much and how long a person is in a contagious condition to the evolution of the disease in the agent.

Upon contagion, the infected individual gradually develops larger and larger levels of virus, in pace with the viral reproduction capabilities in the infected patient. Eventually, a maximum level is reached and the viremic load subsequently declines along with a recovery from infection. This process may be interrupted at any time because of complications, be them virus-based or any other.

We assume therefore that the viremic load is the biological origin of both the severity of illness for an average infected individual and the capability to transmit the virus. In simpler words, the quantity of virus in each individual regulates how ill she/he is and with which efficiency the infection can be passed along.

Symptoms, severity and contagiousness are different from person to person, but they follow an approximate sequence from zero up to a maximum value, subsequently decaying towards zero again. From the day of clear symptom onset we adopt a model for the viremic load, based in early findings [18, 21] from the initial period of the pandemic where individual cases could be traced in detail. We model the viremia from days 5 to 10 using a gamma distribution. The *presymptomatic period* (a period usually of weak symptoms) is modelled in three stages, a first non contagious compartment lasting a day in average, followed by a low contagion compartment, lasting about two days, with the same viremic level than the last day of contagion and finally followed by a compartment with higher contagiousness lasting also about two days.

The duration and distribution of the presymptomatic days, from contagion to symptoms, described in this form is supported by the distribution of times between the appearance of earlier symptoms and the day of diagnostic for the data collected in Argentina (see Figure 2). In fact, the observed mean for the data points is 3.86 ± 0.5 days, plus one day without any symptoms, yielding slightly less than five days before the onset of recognized symptoms and the decision of swabbing.

**Figure 1:**
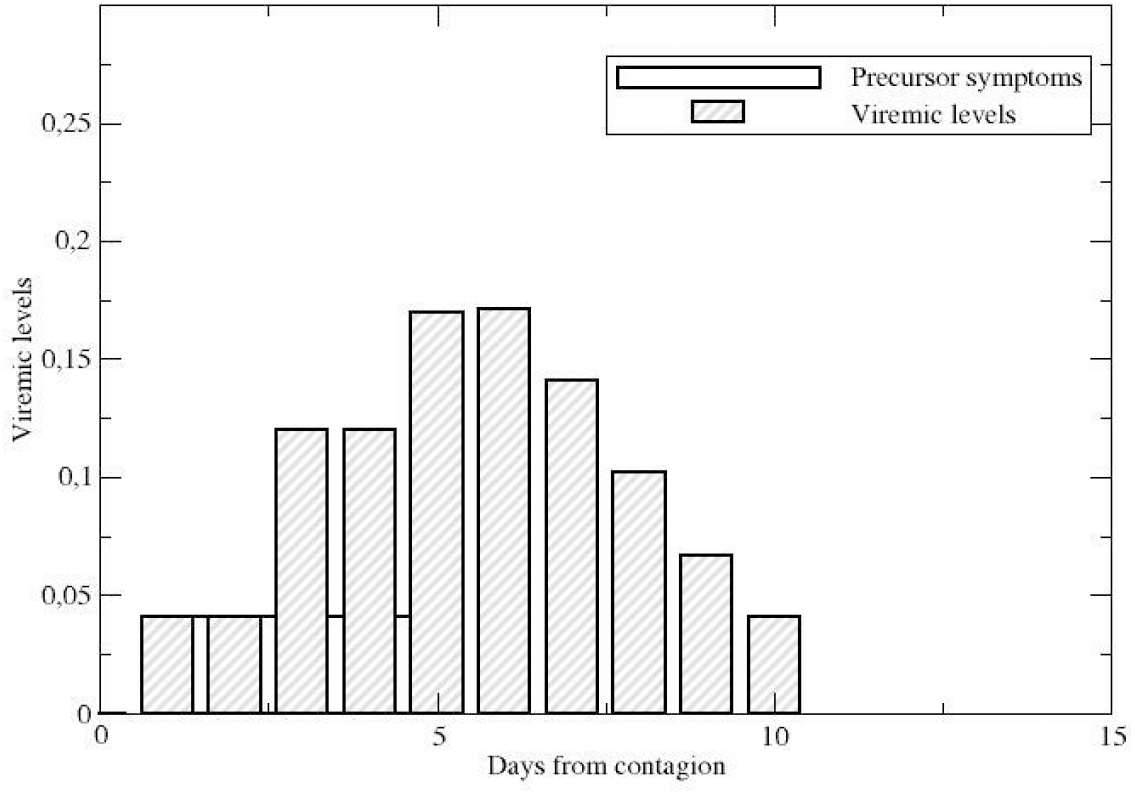
Viremic levels displaying a two-day stage with low virus levels, followed by a stage with higher viremic levels but still no detection symptoms. Bars represent days, not stages.

**Figure 2:**
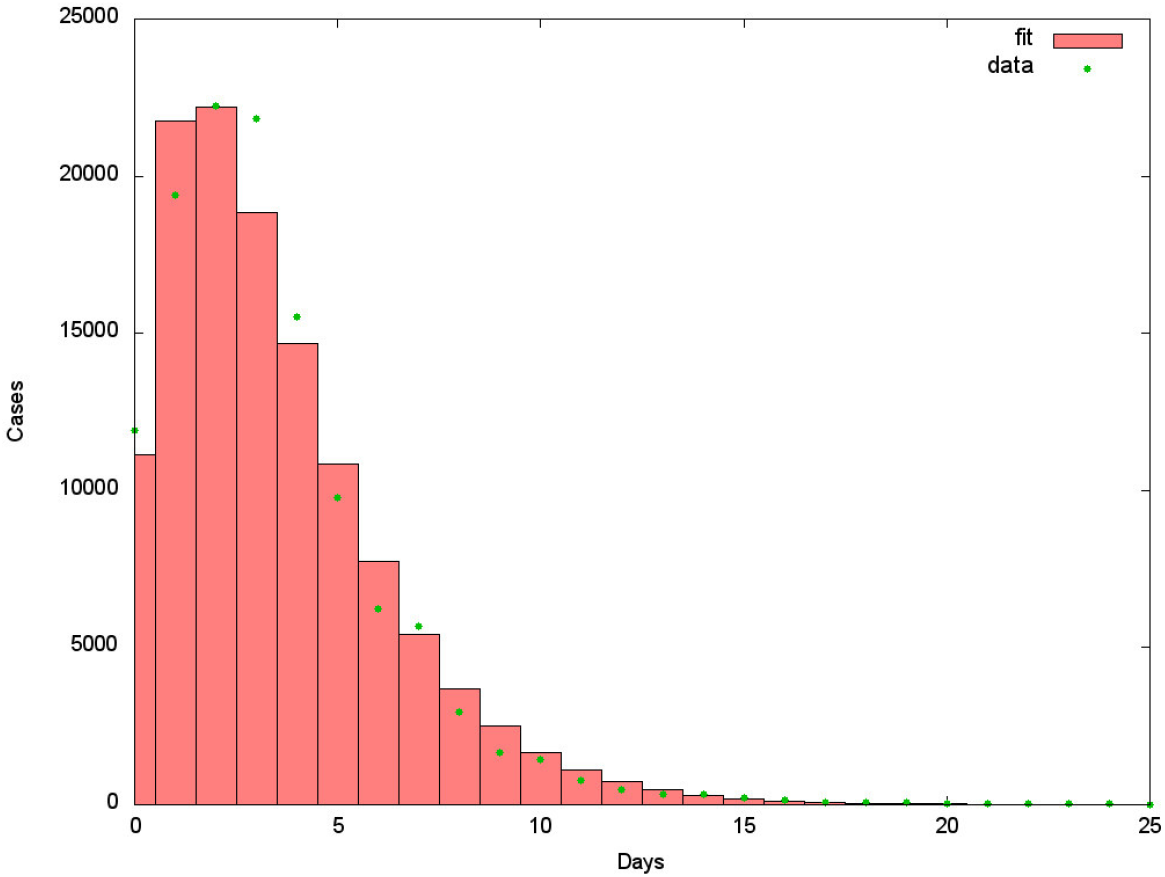
Distribution of time between initial symptoms and swabbing calculated using 121223 entries corresponding to the cases during the month of August 2020 (dots) and curve fit *y*(*x*) = 421627((histogram). The observed average time in Argentina is of ∼3.86 ± 0.5 days. The fitted curve is the composition of two exponentially distributed stages, 2.10 and 1.86 days long in average. It is important to understand that the data reflects not only a biological matter but it is also affected by public health decisions, the information of the population and self diagnosing of the patient concerning the initial symptoms. As such, the statistical error is not the most relevant error. At the beginning of the outbreak the average time was longer than 5 days.

After the presymptomatic period, symptoms usually appear clearly until they gradually decline. We assume the symptomatic compartments to last in average one day each, with viremic levels as in the final part of Fig. 1.

For the sake of dealing with a pandemic, symptoms in themselves are only an ingredient. They facilitate the possibility of detecting infected patients, especially when the pandemic constrains the sanitary authorities to keep a passive attitude. In any case, the appearance of symptoms on each individual depends not only on the viremic load but also on the individual condition of each patient.

On the other hand, regardless of symptoms (if and when they appear), the two processes driving the evolution of the pandemic are *contagiousness* and *detection*. The first one is of course mandatory since there is no pandemic without infections. Both these processes have a social component and a biological component. The biological component was discussed above: We assume both the probability of detection and the probability of contagion to be proportional to the viremic levels of infected individuals, modelled according to Figures 1 and 2.This assumption rest partly on the observations in [18, 21]. However, recent work ([22], appeared after this submission) suggests that there are other mechanisms operating as well, depending on the patient’s response to the infection and deserving a deeper analysis. The modelling profile is summarized in Table 2. The social component reflects the ability of the sanitary authorities to enforce measures in order to (a) detect infected individuals and reduce the chances of contagion (by isolation, hospitalisation, etc.) and (b) effectively influence social behaviour, aiming to reduce the chances that infected, undetected individuals may transmit the disease.

**Table 2:** Normalized viremic levels describing an average evolution of the disease. The probability of contagion is assumed to be proportional to the viremic levels along the different stages. The levels enter the modelling of the probability of detection as well (along with other important influences such as sanitary policies). The levels are normalized so that Σ_*i*_ *V*_*i*_*t*_*i*_ = 1, where *t*_*i*_ is duration and *V*_*i*_viremic levels.

| Stage | Duration (Days) | Normalized viremic level |
| --- | --- | --- |
| 0 | 1 | 0 |
| 1 | 2.10 | 0.0406 |
| 2 | 1.86 | 0.1201 |
| 3 | 1 | 0.1695 |
| 4 | 1 | 0.1713 |
| 5 | 1 | 0.1413 |
| 6 | 1 | 0.1019 |
| 7 | 1 | 0.0667 |
| 8 | 1 | 0.0406 |

#### 2.1.3 About slums

Slum areas have a very specific social structure. They have high population density and intense poverty, resulting in homes shared by several generations (sometimes with just one bedroom), larger number of homeless people in comparison to the rest of city, precarious services (water, electricity, sewage, …), as well as strong internal ties and social organizations such as community run food assistance and other internal solidarity networks. “Stay at home” policies cannot be sustained, to the point that sometimes the whole slum area has been locked allowing for its internal life to continue undisturbed. The contact structure for individuals in slums is more homogeneous and with larger contact rate than the surrounding cities.

#### 2.1.4 The detection of cases as a function of the surveillance protocol

The decision of admitting a case as a probable case of COVID-19 depends not only on the biological/health condition of the case (i.e., the viremic level, presence of symptoms, etc.) but also on the expectations of the health services, HS, as we have discussed in Section 2.1.1 and Appendix A. Since the chances for a contagious person to produce new cases depends on a-priori expectations, the expectations change the removal rate of contagious people (e.g., by isolating the person). Furthermore, the condition of being suspected a-priori is mostly hereditary. The suspicion increases the probability of detection and the detection of a case makes those infected by the case more likely to be detected. Let us call *T, traceable*, those with larger probabilities of detection a-priori, and *U, untraceable*, those with smaller probabilities of detection. Let us further consider the limit situation where all *T* are traced and detected with certainty and no *U* is ever detected. Such an idealized, limit situation will result in two independent epidemics, for no *T* can ever produce a *U* case and reciprocally, no *U* can produce a *T* case. No real situation is expected to reach this limit case, hence, in a more accurate description *U* cases are detected with lesser probability and later than *T* cases. Also, some *T* may escape tracing and detection when still contagious. The inheritance of the tracing classes is then imperfect and there is only one, mixed-type, epidemic. We represent this situation by a probability table (written in matrix form)

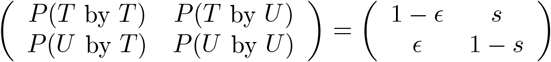

The probabilities, *P* (*X* by *Y*) indicate the probability for a susceptible person infected by a contagious case of type *Y* of becoming a case of type *X* assuming it was effectively infected. The non-negative quantities *ϵ, s* are not new parameters since we have to satisfy that *P* (*T* by *T*) is exactly equal to the probability of a *T* case being effectively detected. The same can be said of *P* (*U* by *U*) with respect to the undetectable cases *U*.

Since all health systems have limited resources and suffer different epidemic impacts, different strategies are likely to appear. One of the goals of this manuscript is to explore the impact of different strategies on the (local) evolution of the pandemic. Schematically, we will consider three scenarios, labelled *passive, intermediate* and *active* representing different policies for the detection process.

In the *passive* policy intervention starts when and if the symptoms are clear. The intensity of the perceived symptoms is assumed to be, on average, proportional to the viremic state. A distinction is made between the *T* and *U*, being the HS’s more prone to act for the *T* group than for the *U* group. The passive policy represents the policies adopted during the early days of the pandemic (mid-February to mid-March 2020 in Europe), where the HS focused attention on imported cases (travellers) and their contacts. The *intermediate* policy reflects the situation in which the HS become aware of the problem of presymptomatic contagious cases, and begin to track oligosymptomatic cases in the *T* group (contacts of known cases). At the same time, it has been observed a lowering on the requirements, in terms of a lesser number and a larger set of symptoms, required for sanitary intervention (isolation). The *active* intervention consists in one of two possibilities: either the *T* class is substantially enlarged by including in it the contacts of contacts, as was done in Italy or by dropping the distinction between *U* and *T* and acting (or strongly exhorting to individual action) on cases presenting any symptom compatible with COVID-19, no matter how weak, as it was the public advise of e.g., the Swedish HS^13^. We will only model the second case.

### 2.2 Mathematical/Computational support

The general approach is based on a Markov-jump process following the setup of the Feller-Kendall [3, 4] algorithm. The compartments *X*_*i*_, *i* = 1, …, *N* involved in the process are the different classes of individuals taken into account (to be described below) and the stochastic dynamics evolves by expressing the number of individuals on each compartment as a function of time. Transitions between compartments are given by Markov jumps triggered by different *events* and characterized by an event probability rate *W*_*α*_(*X*), *α* = 1, …, *E*. The relation between events, compartments (populations) and stochastic dynamics is given by

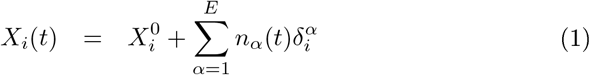

where 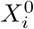 is the initial condition for compartment *i, n*_*α*_(*t*) indicates the number of occurrences of event *α* up to time *t* and 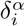 is an integer indicating how each occurrence of event *α* modifies the population in compartment *i*. For the present problem, *δ* will take the values −1, 0, 1, meaning that e.g., one infected individual is removed from the contagious process by isolation, etc. The stochastic dynamics proceeds by establishing the behaviour of *n*_*α*_(*t*).

General properties of Markov jump processes are assumed to hold for this problem, in particular that events are independent of each other (although related indirectly by the dependence of the rates on the populations). These properties add up to the following two results [3, 23, 24, 25]:

1. The waiting time to the next event is exponentially distributed with rate 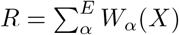.
2. At the occurrence time indicated above, the probability of occurrence for event *α* is 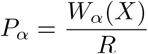.

A realisation of this stochastic dynamical process requires a good knowledge of the probability rates *W*_*α*_ and the computation of one random number (exponentially distributed) for the time of occurrence of the next event and another (uniformly distributed) for selecting the event happening at that time. Upon occurrence of each event, populations and consequently transition rates are updated according to eq.(1). Random numbers were generated with the *Double precision SIMD-oriented Fast Mersenne Twister (dSFMT)* algorithm [26], implemented in *C*.

### 2.3 Details

The algorithm is implemented as a C-program, fully available from GitHub. The compartmental structure is as follows (see Fig. 3):

**Figure 3:**
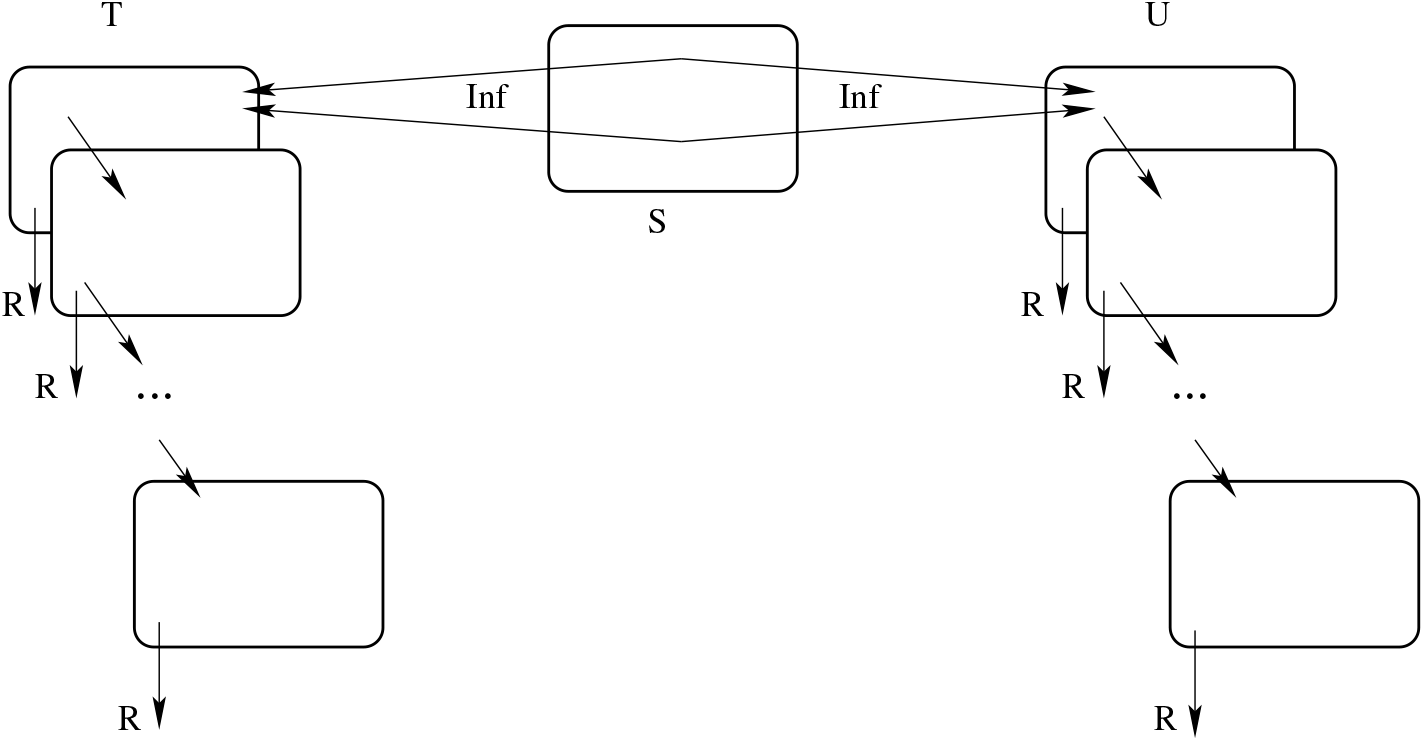
Schematic view of compartments and events.

There exist three classes of compartments, namely *susceptible S, traceable infected T* and *untraceable infected U*. Infected individuals belong in several sub-compartments describing the degree of evolution of their disease (or rather their infective period). At each stage, they may proceed in the disease to the next stage of infection (diagonal arrows) or be removed from the system by any reasonable means, e.g., by being detected and isolated by the HS, by self-isolation, hospitalisation, etc. (vertical arrows labelled *R*), thus ceasing in all such situations to be a source of contagion. Infection may proceed either by contact of *T* or *U* individuals with an *S* individual, or by “importing” the infection from outside the system in consideration.

What regards infection by contact, the tracing of infections is usually not complete, for various reasons. To take this fact into account, we assume that a portion of infections by *T* individuals (of size *ϵ* < 1 in Table 3) may remain undetected and also that a portion of infections by *U* individuals (of size *s* < 1 in Table 3) will eventually become detected. The quantities *s* and *ϵ* were discussed in Section 2.1.4 and will be further specified below. Two additional uniformly distributed random numbers *r*_1_ and *r*_2_ (in [0, 1]) are computed to decide these outcomes (double arrows from *S* in Fig. 3), representing the probability pairs {1 −*ϵ, ϵ*} and {*s*, 1− *s*} respectively for *T* and *U* infected individuals.

**Table 3:**
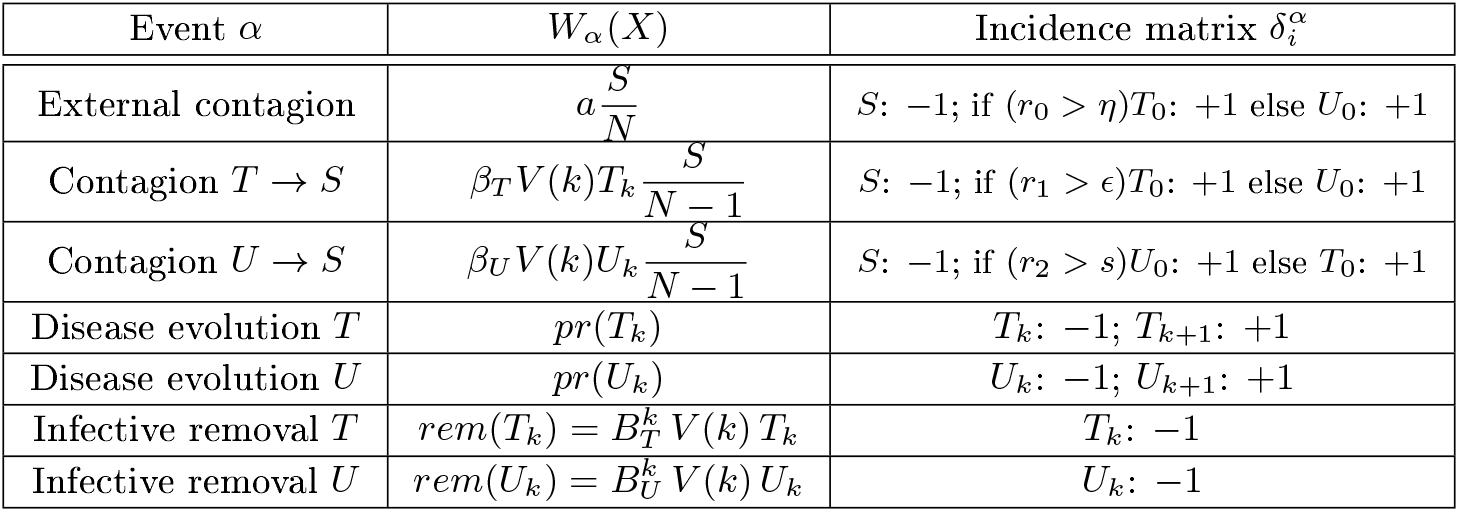
Transition rates and incidence matrix. *V*(*k*) indicate the viremic levels at stage *k. r*_1_, *r*_2_ are uniformly distributed random numbers in [0, 1]. Other quantities are described in the text.

| Event $\alpha$ | $W_\alpha(X)$ | Incidence matrix $\delta_i^\alpha$ |
| --- | --- | --- |
| External contagion | $a \frac{S}{N}$ | $S: -1; \text{ if } (r_0 > \eta) T_0: +1 \text{ else } U_0: +1$ |
| Contagion $T \rightarrow S$ | $\beta_T V(k) T_k \frac{S}{N-1}$ | $S: -1; \text{ if } (r_1 > \epsilon) T_0: +1 \text{ else } U_0: +1$ |
| Contagion $U \rightarrow S$ | $\beta_U V(k) U_k \frac{S}{N-1}$ | $S: -1; \text{ if } (r_2 > s) U_0: +1 \text{ else } T_0: +1$ |
| Disease evolution $T$ | $pr(T_k)$ | $T_k: -1; T_{k+1}: +1$ |
| Disease evolution $U$ | $pr(U_k)$ | $U_k: -1; U_{k+1}: +1$ |
| Infective removal $T$ | $rem(T_k) = B_T^k V(k) T_k$ | $T_k: -1$ |
| Infective removal $U$ | $rem(U_k) = B_U^k V(k) U_k$ | $U_k: -1$ |

For the “imported” infections taking place outside the system, the uniform random number *r*_0_ distributes the resulting infected individuals among *T* and *U* with proportions {1 − *η, η*}.

#### 2.3.1 Rates and actions

In Table 3 we describe the expressions adopted for the different rates and their action on the population (i.e., the nonzero values of the *incidence matrix* 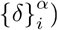). Considering the nature of the available data, the time-unit is (*day*)^*−*1^, i.e., transition rates are given per day. In the table, *N* is the size of the population, typically a neighbourhood or other region that can be safely assumed to behave homogeneously (basically, that any individual may in principle meet any other individual; a natural assumption for working places, schools, etc.). Initial conditions for all simulations is that most individuals (*N* − *c*) are in compartment *S* while the remaining *c* are in *T*_0_ or *U*_0_ (we assume *c* is typically around 2 for *N* up to a few thousands). The “import” rate 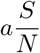 describes infected individuals undergoing contagion outside the system. We include in this event the possibility of travellers bearing the infection when returning to the system after a temporary absence, a group that has been important in global scale to transfer the disease across continents, but is comparatively small for such stable communities as those we consider.

The evolution of the illness is given by stages *T*_*k*_, *k* = 0, …, *K* − 1 and similarly for *U*) describing the viremic level *V* (*k*) at each stage (Figure 1 and Table 2). In this work, *K* = 9. *pr* describes the rate of passage to the next viremic stage i.e., (*pr*)^*−*1^ is the average permanence of an individual on each stage (second column in table 2). The factor *βV* (*k*) describes the contagion rate between a susceptible and an infected individual. In principle, *β*_*T*_ and *β*_*U*_ may be different but we have not explored that possibility. Since the viremic levels are normalized, the weighted infective period is one and the factor *β* corresponds to the basic reproductive number *R*_0_ of the SIR model. The approximate value of 2.5 was taken from early reports from China [27] and contrasted against data from the initial days of the first outbreak in slums in Argentina (Barrio Padre Mujica, also known as Villa 31 in Retiro, a neighbourhood of Buenos Aires city)^14^.

The constant *ϵ* indicates what portion of the individuals infected by a *T* will not be detected by the health services during the contagion phase of that case, while similarly *s* indicates what portion of the individuals infected by a *U* will eventually be detected by the control procedures. Similarly, the constant *η* describes the distribution of imported cases among *T* and *U* compartments. Since *η* is largely unknown we will only consider the extreme cases. We have *η* = 0 usually associated to long distance travels to/from regions of viral circulation. For the case of slums, casual contagion within the same city but in e.g., different neighbourhood is expected to be the most frequent case, hence we adopt *η* = *s*.

Finally, *rem* is the rate of removal of an individual from stage *k* out of the contagion chain. This rate also depends both on the viremic level and on the HS strategies (giving different choices for the factors *d*_*T*_ (*k*), *d*_*U*_ (*k*)). This part of the model will be described in detail in the next Subsection.

The data usually discussed in the news and websites is the number of confirmed COVID-19 cases. In the present model this data is represented by the total number *D* of detected individuals, i.e., the outcome of all removal events within the infective period. The model provides an estimate of the silent cases, i.e., infected individuals of which the HS has no records. In the model these nondetected infected individuals *ND* are given by the identity *N* = *S* + *D* + *ND*, where *N* is the population size and *S* the number of susceptible individuals.

The outcome of the model is presented by computing a few realisations (typically 100) of the Feller-Kendall algorithm. No matter how parameters are chosen, there exists a non-zero probability of early disease extinction (as it is in any Markov jump process), particularly when the onset of the epidemics contains very few infected individuals (1 or 2 on a population of a few thousands). The model allows for ruling out early extinctions, considering that the epidemics that are tracked, and concern us, are those that avoid early extinction and come to be noticeable.

The actual evolution of the pandemic is intrinsically stochastic. Borrowing from the modelling language, there is only one “realisation” of the real process, namely the one we are currently experiencing. There is no “second run”, although many weakly coupled contagion chains may be running simultaneously within e.g., a larger city.

With this in mind, we stress that the averaging of realisations is not a substitute for the real process. It has a limited value, in that it highlights features that are recurrent, while it smears out what is less frequent. Moreover, no realisation of the stochastic process is “more true” than any other. Predictions based only on the averaging of realisations may serve as a clue about what to do, but policy decisions should take into account the whole picture.

#### 2.3.2 Contagion, removal and HS policies

We consider in detail the mechanisms of contagion and removal, as well as their relation with both the evolution of the disease in the infective individual and the HS policies.

Contagion within the system is taken to be strictly proportional to the viremic levels *V* (*k*). The proportionality constants *β*_*T*,_ *β*_*U*_ may vary according to social strategies and attitudes.

The eventual removal of an infected individual in the model is governed by the competition between two mutually exclusive events. Either the individuals evolve to the next stage in the viremic levels (i.e., they are still infected and capable of contagion) or they are removed from the contagion chain for whatever reason (detection, isolation, full recovery or death). At stage *k*, the probabilities 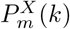 of moving to the next stage in the contagion chain and 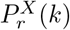 of being removed from the chain for an individual of class *X* = {*T, U*}, can be described (in the notation of Table 3) as:

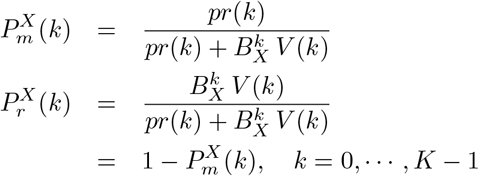

where 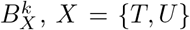, model the HS policy adopted. In the present implementations, the factors 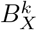 are set to zero for an initial subset [0, …, *k*_0_ − 1] of stages (*k*_0_ ≥ 1) and take the same positive value *B*_*X*_ for the remaining stages, *k* ∈ [*k*_0_, *K* − 1]. *B*_*X*_ relates to the probability of *X* being effectively detected (named 1 −*ϵ* and *s* in Section 2.1.4) through Eqs.2 below.

At the final stage, *K* − 1, the overall action of both competing events is a removal from the contagion chain. Individuals that have not been removed at any previous stage, have effectively participated in the contagion chain during all of their contagious period. These individuals were not detected by the HS policies while they still were active in the contagion chain. The overall probability of detection can be computed as follows. Let 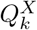 be the probability of removal up to and including stage *k* for infected individuals of class *X* = {*D, U*}. Set further,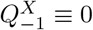. For any stage *i*,

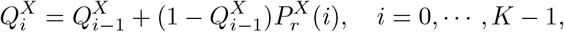

which can be restated as 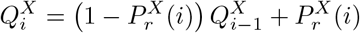. The total probability of removal during the infective period is 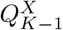, while the probability of being *detected* at some point during the infective period for individuals in class *T, U* is given by

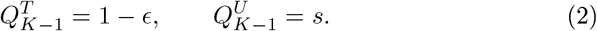

Note that 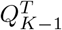 and 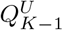 are rational functions, the ratio of two polynomials of degree *K* − *k*_0_ in *B*_*X*_.

Eq. 2 relates the value of the constants *B*_*T*_, *B*_*U*_ for the different HS policies with the probability of detection. The differences in dealing with *T* and *U* infected individuals follows from the differences between 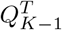 and 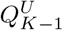, being the HS’s more prone to act for the *T* group than for the *U* group. We distinguish three main policies:

#### Passive policy

Intervention on the *U* class concerns only severe cases (e.g., requiring hospitalisation) in a situation where the viremic levels of the patient are comparatively high. In the model, intervention for the *U* class starts at *k*_0_ = 3 (stage 3 in 2).

#### Intermediate policy

The conditions required for sanitary intervention (isolation) in the *U* group are broadened in terms of a lesser number and a larger set of symptoms and possibly intervention at an earlier stage. Active (preventive) intervention, as in contact tracing, starting at stage *k*_0_ = 0 (or 1), is implemented for the *T* group. It reflects a situation in which the HS become aware of the problem of presymptomatic contagious cases, and begin to track oligosymptomatic cases in the *T* group (contacts of known cases).

#### Active policy

No distinction is made between *U* and *T* regarding actions of the HS. One symptom is enough to trigger sanitary actions. Interventions start at stage *k*_0_ = 0 (or 1).

#### Simulation scenarios

In the next section we discuss a few scenarios based on these policies, relating to data from Table 1. We consider three detection efforts, that we may call *Low, Medium* and *High* (*L, M, H*) for each scenario. We identify the *T* group with health workers, who for practical reasons were better monitored than other individuals. A first scenario, labelled *I* (for “Ideal”) corresponds to the active policy, with three different effort levels, represented by *B*_*X*_ such that the detection probability 1 − ϵ= *s* takes the values 0.21, 0.61 and 0.79. The latter corresponds to the fraction of confirmed cases among health workers displaying one symptom of the extended list of Section 2.1.1. However, registration of symptoms was optional. Therefore, 0.79 is only a crude lower bound to the ability of detecting cases among health workers (which are subject in part to routine testing). A second scenario of intermediate character, labelled *F* + (for “fever plus other”), corresponds to the same detection probabilities as above for the *T* group, whereas for the *U* group the detection probabilities *s* are set to 0.10, 0.28 and 0.36. The latter corresponds roughly to the proportion of confirmed cases among health workers displaying fever plus another symptom in Table 1. The third scenario, labelled *H* (for “hospital”) is still unaltered for the *T* group relative to the previous two, while the *U* group is subject to the Passive policy (thus assuming that only highly viremic cases have a chance of being detected, and only from stage 3), with detection probabilities *s* set to 0.02, 0.06 and 0.08. The latter corresponds roughly to the proportion of confirmed health worker cases that were hospitalized in Table 1. Hence, the *High* intensity level of the three scenarios relate to detection policies adopted by HS’s at different periods of time. The lower estimate for the HS actions is roughly 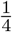 of the upper estimate and the intermediate estimate was taken to obtain an intermediate level of detection. In the unfortunate situations where the epidemic is out of control, the effectiveness of the HS measures could be even lower.

Unless otherwise stated, all simulations are performed with 5000 individuals, of which two are initially contagious in the *T* compartment (it makes only imperceptible difference to set the initial contagion in the *T* group or the *U* group), while the contagion rate is set to *β* = 2.5 and there is a small rate of external contagion (*ext* = 0.002).

A list with the parameter values used in different scenarios can be found in Table 4. Other necessary input data for running the simulations is: number of realisations (usually 100), length of simulation in days, initial condition for populations *S, T, U* (usually 4998, 2, 0), random number seed, flag to discard “early” extinctions (positive integer) and maximal duration to be considered “early” (usually 19 days).

**Table 4:** Parameter values used in various simulation scenarios, in the variants low, medium and high respectively. *B* is called *det* in the code and *k*_0_ is called *delay*. Under the present normalization, the contagion rate *β* corresponds to the basic reproduction number *R*_0_ of the SIR model.

| Name | Ideal $I$ | Fever+ $F+$ | Hospital $H$ |
| --- | --- | --- | --- |
| $a$ | 0.002 | 0.002 | 0.002 |
| $\beta_T = \beta_U$ | 2.5 | 2.5 | 2.5 |
| $B_T$ | 0.24,1.00,1.76 | 0.24,1.00,1.76 | 0.24,1.00,1.76 |
| $k_{0T}$ | 1,1,1 | 1,1,1 | 1,1,1 |
| $B_U$ | 0.24,1.00,1.76 | 0.10,0.34,0.46 | 0.03,0.09,0.12 |
| $k_{0U}$ | 1,1,1 | 1,1,1 | 3,3,3 |

## 3 Results

### 3.1 General Results

The following results follow from the structure of the model. There is essentially nothing left to prove, just following the construction in 2.3.2.

#### Lemma 1.

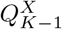 *is a monotonically increasing function of B*_*X*_.

In modelling language, *B*_*X*_ senses the efficiency of the detection process.

#### Lemma 2.

*For fixed B*_*X*_, 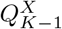 *is decreasing with increasing k*_0_.

A late start stage for the detection process can be interpreted as a HS policy that is only capable of taking care of seriously ill cases, with highly developed viremic levels. The more stages an individual passes without any policy action, the lower the overall chances of detection within the infective period.

### 3.2 Simulations

Simulation results allow us to compare the outcomes of different policies on an equal footing.

#### 3.2.1 Spread

Before considering averaged results let us sense the spread of outcomes from different realisations of the process. In Figure 4 we show the fraction of susceptible individuals as a function of time for 100 realisations of the stochastic process in two different configurations.

**Figure 4:**
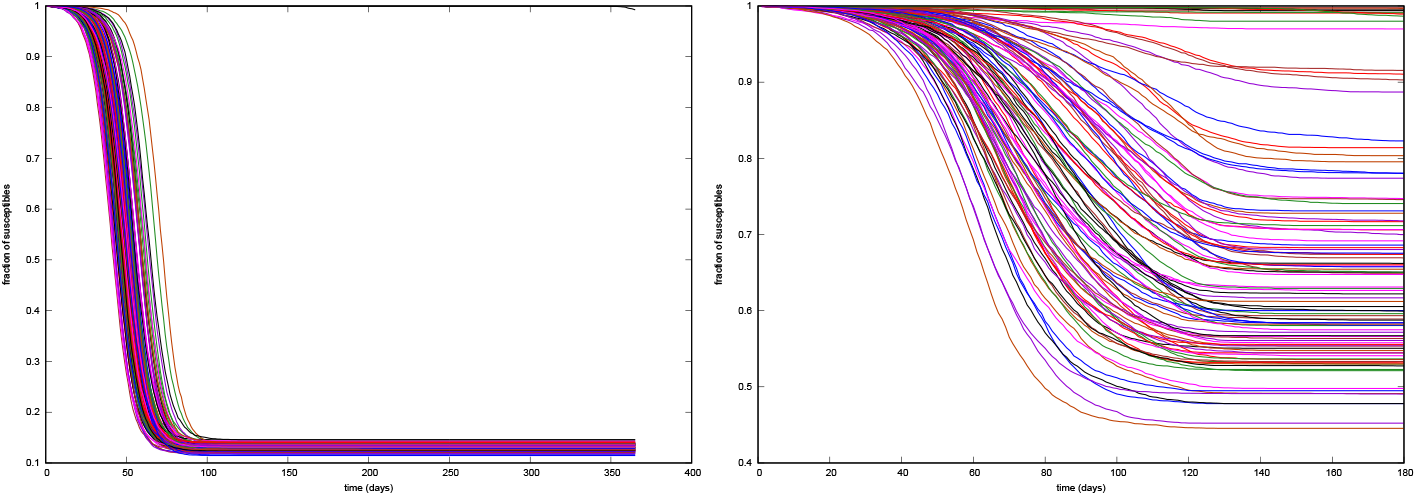
Fraction of susceptible individuals for 100 realisations. Left: Low, constant detection. Right: Increasingly sharp detection

The left panel corresponds to an situation where the probability of detection while still contagious is 21% for the *T*-group and 9% for the *U*-group, with *β* = 2.5. All outcomes display a sharp fall in the number of susceptible individuals. Notice however the spread in time: The fastest and slowest realisations differ in about 40 days, corresponding to 100% at the 0.5 level. The right panel corresponds to a weaker contagion situation (*β* = 1.75), where the probability of *T*-detection increases every 60 days, from 0.6 through 0.71 up to 0.79 (all detections starting on stage 1), while the *U*-detection goes from 0.07 through 0.58 (with detection starting on stage 3) up to 0.79, with detection starting on stage 1. Notice here the spread in the outcome. While some realisations display almost no variation in the fraction of susceptible individuals, some others achieve a fall of over 50%. It is worth to keep in mind that the policy of progressively increasing the detection effort has been the rule in practical cases.

#### 3.2.2 Initial growth

As in most models with homogeneous contact, the initial growth of the epidemic outbreak is almost exponential and this regime lasts for about two months in the present simulations with about 5000 initial susceptible individuals. However, it is worth to indicate that the growth exponent of infected cases and that of detected cases is not the same, being the latter smaller than the former, specially in less effective regimes as *H* and *F* +. As a consequence, basic reproductive numbers inferred from the early development of the pandemic that had assumed that detected cases are roughly proportional to the actual cases underestimate the growth rate. See Figure 5. Note that the gap between growth rates is larger for the lower detection effort as compared with the higher (red/green vs blue/magenta pairs).

**Figure 5:**
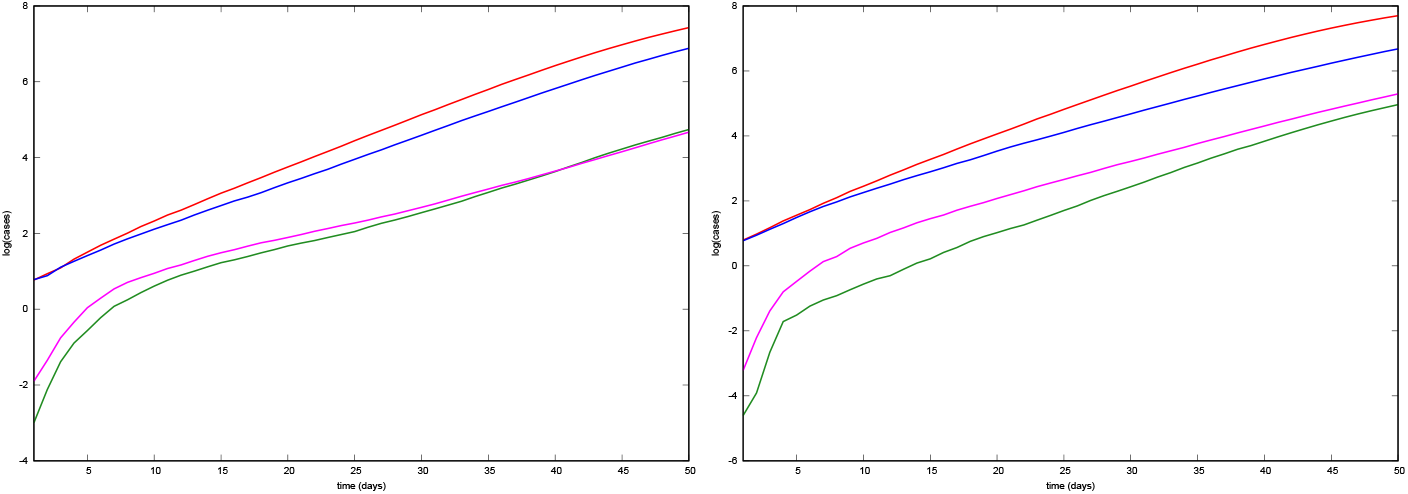
Average growth rates for 100 realisations at the beginning of the epidemics. Left: *H* simulations. Red (log(*N*−*S*)) and green (log *D*) curves correspond to Medium effort while Blue (log(*N*−*S*)) and magenta (log *D*) curves correspond to High effort. Right: *F* +. Red (log(*N*−*S*)) and green (log *D*) curves correspond to Low detection effort while Blue (log(*N*−*S*)) and magenta (log *D*) curves correspond to Medium detection level.

#### 3.2.3 Undetected/detected ratios

The ratio between total cases and detected cases of COVID-19 has been the subject of several works. In particular Malani et al. [28] and Muñoz et al. [29] address the situation in slums, reporting ratios of 10 : 1 [28] and 5 : 1 [29] ^15^. The latter study was performed at least one month after “most cases” occurred, although with the outbreak still running. The majority of the registered cases had occurred before 6 June, when the tracking method in use was of type *F* +. After 6 June, the tracking sharpened to “any two symptoms”, a medium form of *I*. We show averaged ratios in Figure 6 but it is worth to keep in mind that there are usually large fluctuations present. The figure shows that in all situations there is a tendency to a sharp increment of the ratio at the beginning of the outbreak followed by a maximum level and subsequently a monotonic decrement.

**Figure 6:**
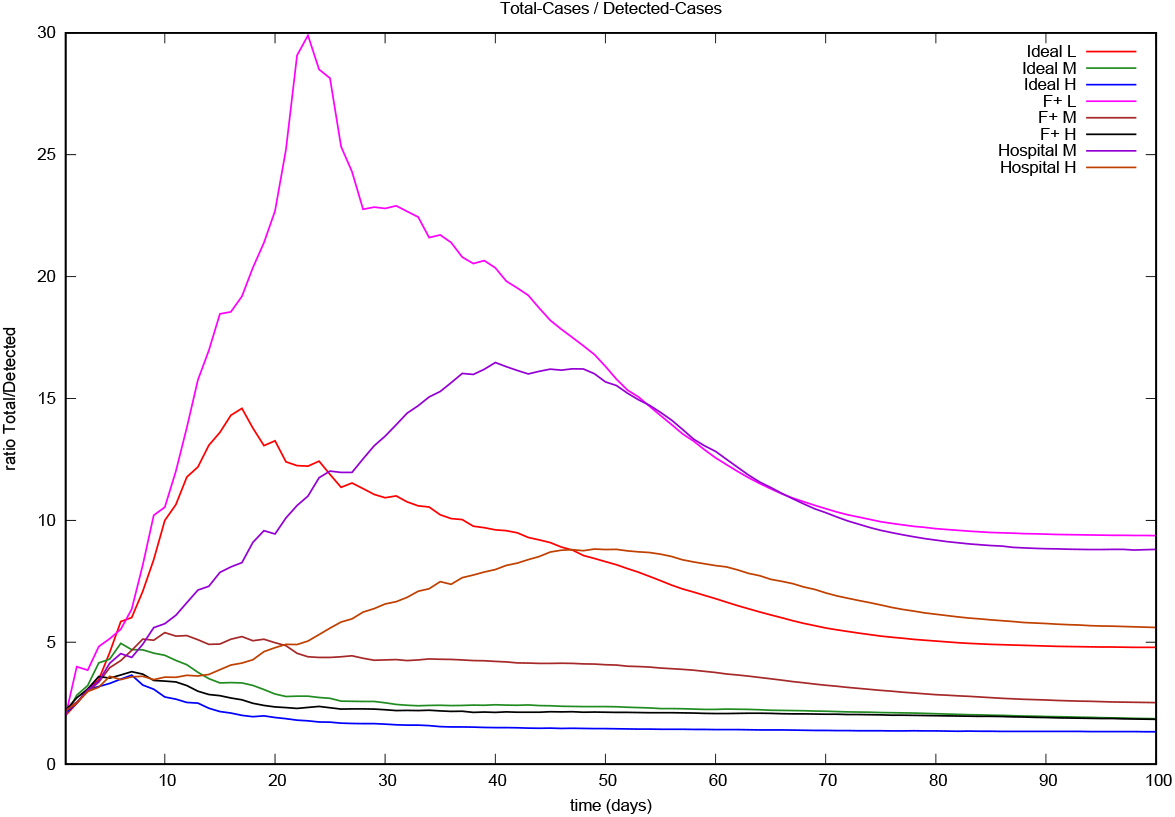
Average ratio of total cases to detected cases (100 realisations) under eight different control measures implemented with moderate efficiency.

#### 3.2.4 Dynamical mechanisms

The most remarkable features present in the simulations are the diverse forms in which the stochasticity and the particularities of the contagious process manifest globally. Despite being seeded with two traceable cases, it is not uncommon (probability larger than 0.01) to observe the outbreak to remain with sporadic cases up to 50 days and only then the recognisable bell-shaped of the daily cases begins. We show one of such cases in Figure 7, left panel. In the centre panel we show a “two waves” outbreak under policy *IH* and a different shape of “two waves” with a long delay between them is shown in the right panel. The difference among realisations suggests that stochastic epidemic outbreaks are not just an “average outbreak” plus noise.

**Figure 7:**
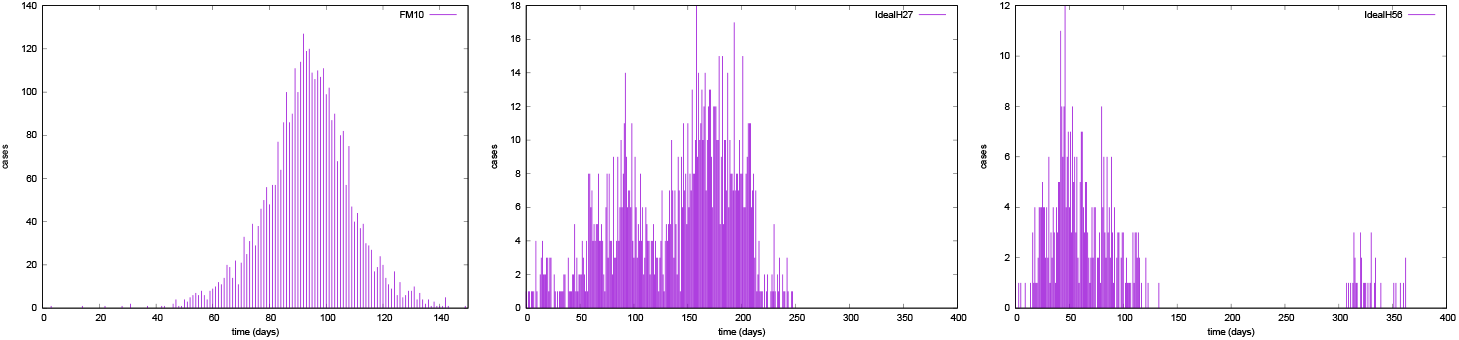
Daily cases. Observable delays in the development of outbreaks for three individual realisations. Left, F+M policy presenting a “long” waiting time until the outbreak develops. Centre, *IH* policy with immediate second wave. Right, *IH* policy with delayed second wave.

As expected, the epidemic size depends strongly on the policy applied. It is interesting to show the transition of the probability distribution as a function of the intensity of the control measures. In Figure 8 we show histograms after 100 simulations for the total number of infections in a population of 5000 individuals. While extinctions of the epidemic with a low number of cases are always possible, they are infrequent in the Low intensity case, they begin to be noticeable in the Medium intensity and are dominant in the High intensity situation. This transition is known as the stochastic equivalent of the transition in deterministic equations when the basic reproductive number moves from above one to below one and has been discussed elsewhere [32, 33, 34].

**Figure 8:**
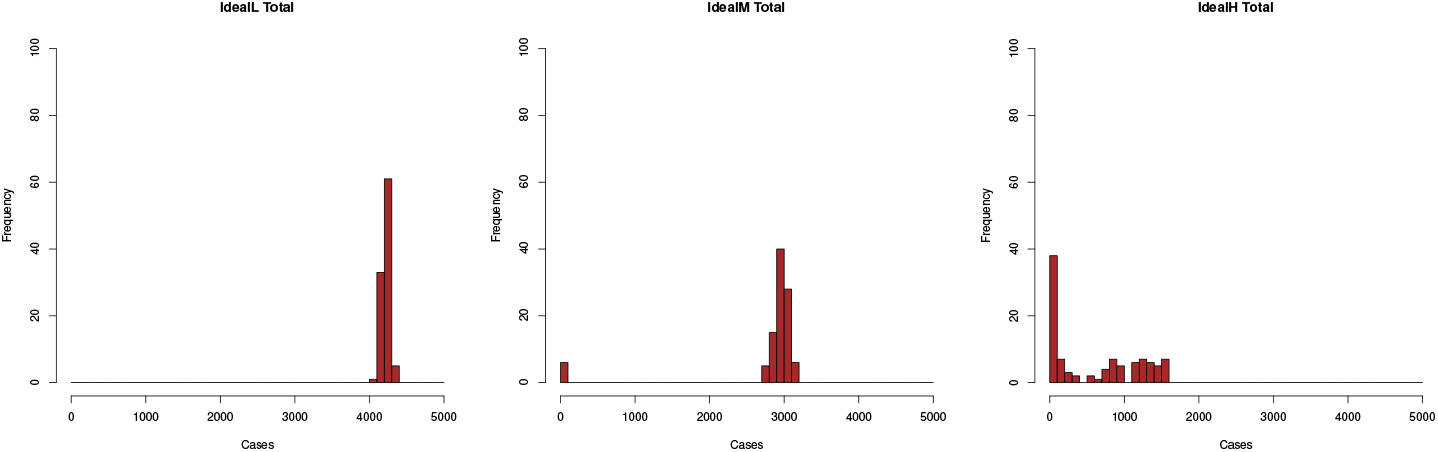
Transition of probability (frequency, %) distribution for the *I* (Ideal) policies as a function of the intensity of control measures. Left, low detection rate. Centre, medium detection rate. Right, high detection rate.

The number of *detected cases*, i.e., people diagnosed as infected with SARS-CoV-2, depend in no trivial form of the detection policy and intensity. We show in Figure 9 that the relation is not monotonic. In general, an increase in detection efficiency from medium to high intensity may result in a decrease in the number of cases detected (policies *F* + and *I*) but also in an increase (*H*). In fact, the design of the model only assures that the *probability* of detection increases with increasing intensity. If the total number of cases is low, the total number of detected cases will also be low, despite a higher detection probability. We can see as well that the efforts made with a passive policy (*H*) produce only little changes in the development of the epidemic.

**Figure 9:**
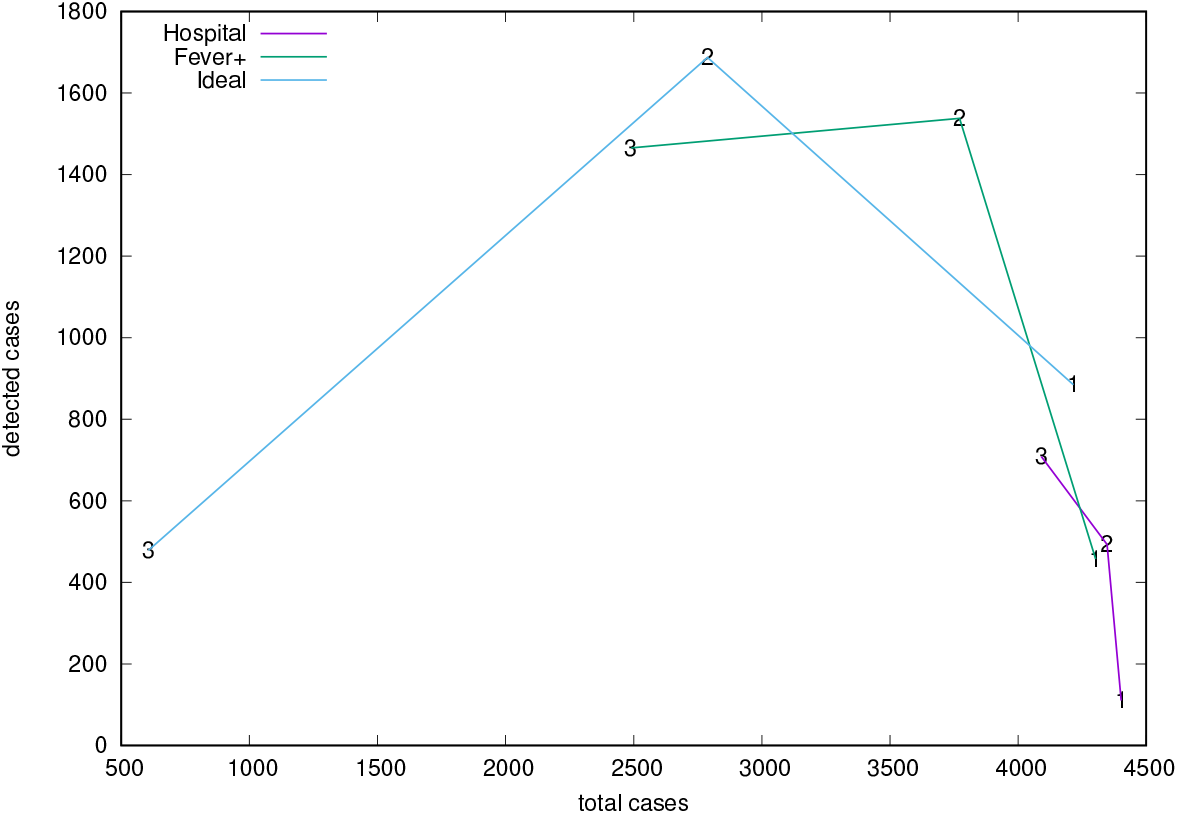
Relation between detected and total cases corresponding to the average over 100 realizations of the policies *H, F* + and *I*, with three intensity levels labelled 1, 2, 3 in the plot and corresponding to Low, Middle and High detection levels.

## 4 Discussion

In the present model biological aspects are intertwined with sanitary policies. These policies are not considered in terms of their desired effects translated as effective parameters of an otherwise free running epidemic but rather mechanistically, changing not only parameter values but the structure of the model as well. By doing so, we allow policies to manifest not only what they were intended for, but also unexpected features. The same can be said with respect to the coupling between intrinsic randomness and dynamics which results not only in the expected daily fluctuations of the outbreak but presents low-frequency fluctuations as well. These low-frequency fluctuations account, by themselves, for the possibility of silent circulation of the virus for prolonged intervals of time as well as for the awakening of extinguished outbreaks due to contagion outside the simulated community. The effective coupling of control measures and intrinsic randomness represents an additional difficulty for any attempt at predicting the evolution of a single/particular epidemic outbreak.

Although it should be clear from the setup adopted in this work, it is worth recalling that all realisations of a stochastic model are on an equal footing. Any of them respond to the process in its own right. A strength of the present approach is the capability of displaying a variety of possible epidemic outcomes. Indeed, Figure 4, Right and 7 show that dramatic differences in epidemic size (for the same stochastic process), “second waves” and late development of outbreaks are not unlikely to occur.

Figures 5 and 6 show that the ratio of undetected to detected cases is not constant in the course of an epidemic and even worse, the growth rate of undetected cases is larger than that of detected cases. Hence, epidemic size is likely to be underestimated when computed through recorded cases. Finally, Figure 8 illustrates how the stochastic outcomes can be translated into probabilities of e.g., having a given epidemic size.

One important goal of this work is to assist in the issue of resource allocation when dealing with a pandemic. HSs throughout the world differ in equipment, logistic capabilities, flexibility, etc., depending on the preexisting policies and infrastructure. The working conditions differ even locally within the same city, as discussed above. Where should resources go? Will low-cost (and lower efficiency) strategies under a longer period of time be preferred to high-cost (and higher efficiency) strategies with a shorter time-span?

In our model we mimic the HS decisions by considering two groups of individuals: Those that are early identified and recognized as potential patients, *T*, and the rest, *U*, of which the HS is initially unaware. We do not deal with financial costs, but we can compare highly-effective and less effective strategies throughout time. Preventive intervention strategies accrue costs in terms of isolating contagious people and testing. The kind of intervention considered in the present work is based upon tracking oligosymptomatic people but not searching for completely asymptomatic cases, thus a good indicator of costs is the total number of cases detected. The best strategy of all those considered in this respect is the *I* strategy with an efficient, *H*, search method, which is able to inhibit the development of outbreaks. The strategy has fewer detected cases and smallest overall size (see Fig. 8, right panel and Figure 10, upper left panel).

**Figure 10:**
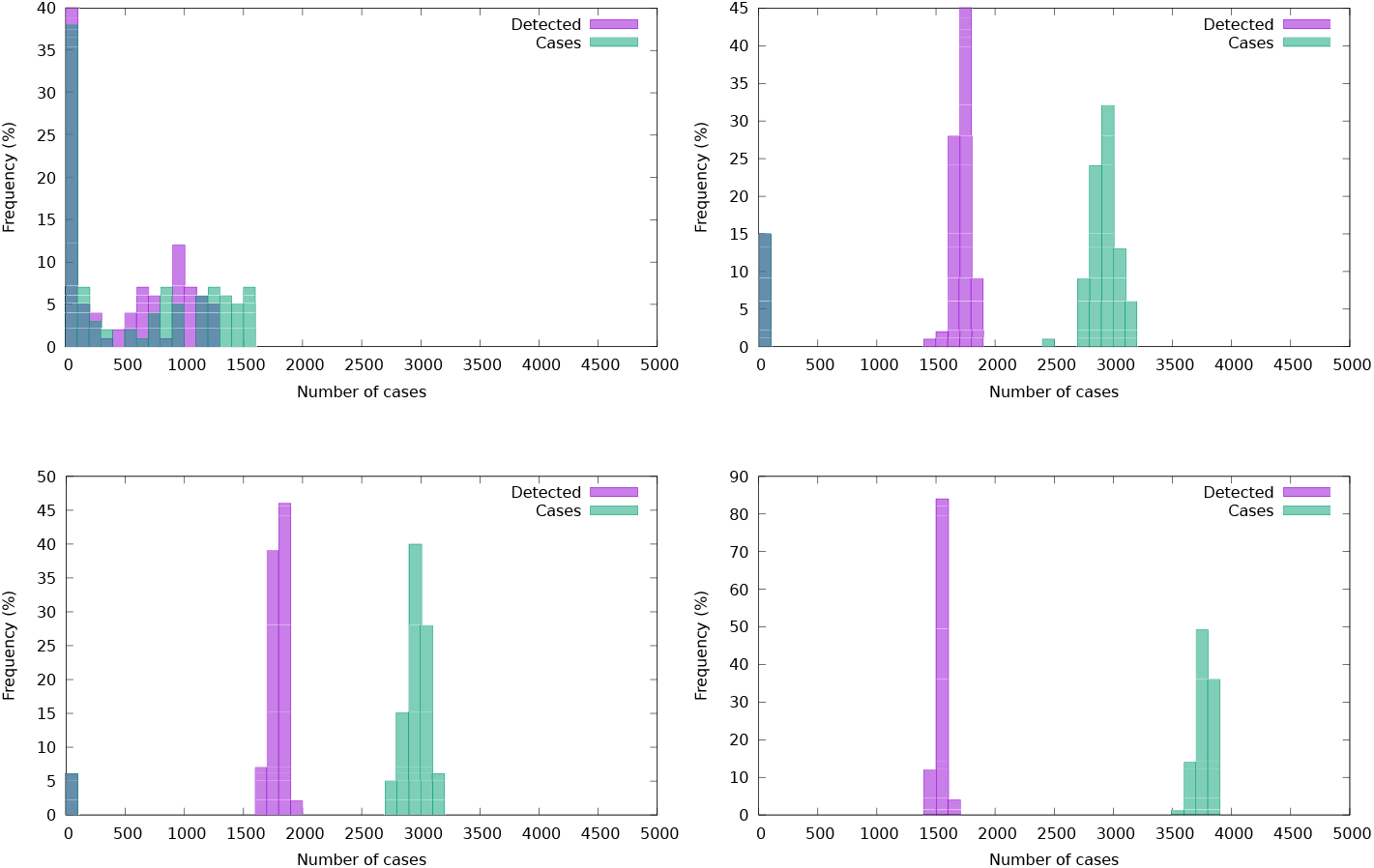
Frequency distribution (%) of detected and total cases. Upper panel, *IH* and *F* + *H*. Lower panel, *IM* and *F* + *M*.

We compare two distributions of detected cases for equal efficiency of detection, see Figure 10. The comparatively few detected in the *I* panel of Figure 10 constitute most of the outbreak, while *F* + adds a larger number of undetected cases. A short side of the *I* policy is that the effective suffocation of an epidemic outbreak in slum areas cannot by itself prevent recurrent late outbreaks triggered by external contagion (see Figure 11). Hence, the alert state of the HS will have to be maintained for longer times. However, the advantages of the *I* strategy under a medium or low detection success, thus having a higher failure rate in avoiding outbreaks, is not so considerable. As it can be seen in the figure, combining a suboptimal policy with a suboptimal tracking (*F* + *M*, lower right panel) is expected to be more cost effective than an optimal policy with suboptimal tracking (*IM*, lower left panel) in terms of detection, although the overall size of the epidemic is expected to be lower in the *I* situation, while the necessary social effort is larger.

**Figure 11:**
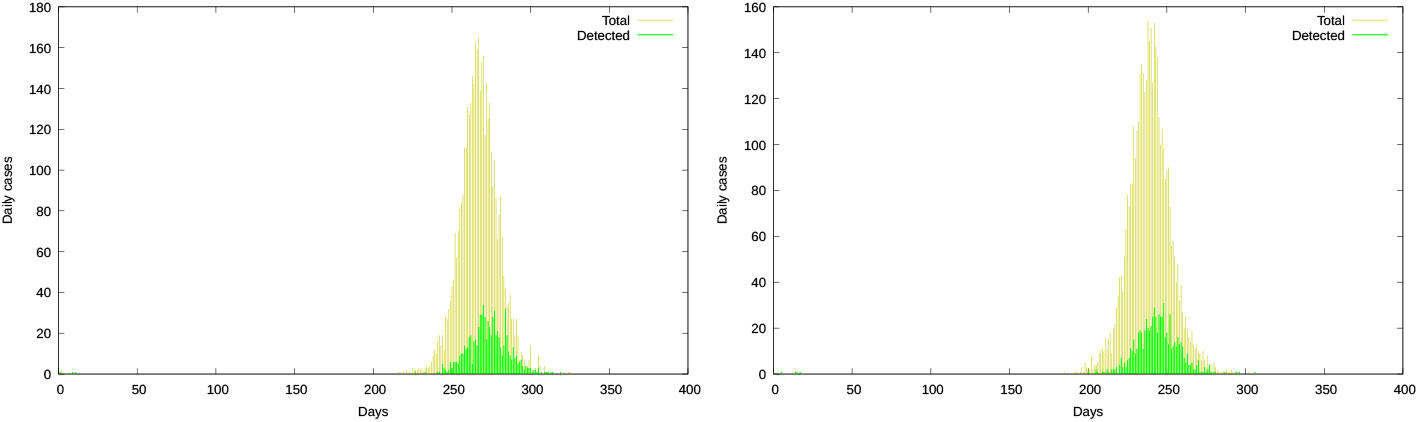
The initial (seeded outbreak) dies out but after an external infection a “delayed” outbreak emerges.

Despite our simulations have been seeded in all cases with two infected people and only epidemic outbreaks that do not get extinguished for 19 days have been considered, some runs do not develop an outbreak and some others produce an outbreak only because of external contagion (from outside of the simulated neighbourhood), an effect than can occur in a completely different time scale as is shown in 11

Also, we illustrate that the “average epidemic” is not enough to grasp the relevant diversity of possible scenarios of real (unique) outbreak dynamics, and that the undetected (mild, unrecognized, presymptomatic, “asymptomatic”) cases are in good proportion the result of public policies coupled to the characteristics of the illness.

### Limitations

As mentioned previously, one of the assumptions of the present model is the homogeneity of contacts through the population. For that reason, it only makes full sense when applied to small communities. The proper path to surpass this constraint is to raise the level of detail, identifying subpopulations with some common property (e.g., age segregation, mobility, local confinement, etc.) that are in weakly mutual interaction. This is a costly approach from the point of view of experimental design, since each new level of detail demands a detailed understanding of the specific interactions. Some effort in this direction has been to identify “superspreaders”, a possibility that recently became interesting. [35]

## 5 Conclusions

The modelling goal of this work was to conceive mechanisms for the interplay of the epidemic disease and the adopted social measures. The epidemic is not just biologically given in terms of e.g., a basic reproductive number or a herd-immunity level that are taken to be virus-specific and independent of social organisation. The COVID-19 pandemic is not a free running epidemic or one addressed with pre-established measures, but rather one where dynamically evolving interventions are the rule. The way in which interventions change the dynamics and the observations (monitoring) of the epidemic must be considered. All too often the analysis of the epidemic by the political components of the HS’s, in practice consider a biologically given epidemic, and interventions that affect its evolution only in the way they were intended. This reasoning leads to false alternatives between herd immunity, vaccines and confinement, with the hidden assumption that social behaviour cannot (or must not) be modified. On the contrary, this work suggests that what we (collectively) do influences the level of risk to which we are exposed. Social behaviour can modify epidemic outcomes [36].

We observe that an increment in the number of daily detected cases does not necessarily imply an improvement on how the epidemic is being managed, nor a worsening of the outbreak. Case-detection cannot be understood separately from the HS policy. Lower detection may be an indicator of success in the proper context. Hence, to translate the statistics for one country to another country is far from straightforward. More locally, the transfer of information from detected (registered) cases to estimated number of cases from seroprevalence studies is not independent of the adopted HS policy and depends as well of the timing with respect to the development of the outbreak.

Randomness plays a substantial role in COVID-19 dynamics, a role that departs from the signal+noise analysis framework. Low frequency, or coherent fluctuations, are relevant at the level of outbreaks in slums and there is no reason to believe the same is not going to be true in larger, heterogeneous, settings. The immediate consequence is that averaging and uncontrolled “approximations” to the average outbreak will be aligned with intuitions but could be misaligned with a reality displaying a largely unpredictable form. The stochastic behaviour is affected as well by the social management of the epidemic, coupling two usually neglected contributions and making prediction of outcomes even more difficult.

The intervention of health authorities had been “from below” in most countries. By “from below” we mean a sequence of interventions going from non-intervention and passing through increasing levels of action until reaching lock downs in desperation. Such an approximation has to be revised, it is an approach that privileges something different than people’s health. If our model is correct, it is possible to control the outbreaks with interventions that target mostly the symptomatic population. Such a method will have to target for isolation of any one presenting a single symptom of those compatible with COVID-19. The cost of more certainties is to lose control of the outbreak, being forced to apply lock downs, thus immobilising the productive forces of the healthy people rather than the comparatively small group that is potentially infected by SARS-CoV-2.

The decision of requiring more symptoms to declare a case as COVID-19 suspect whenever the patient has no identified contact with confirmed cases facilitates the circulation of the virus even when a highly efficient detection protocol is used.

Asymptomatic cases quite often are undetected cases as well, but the reverse is not true. Classifying or referring to undetected patients as asymptomatic can be viewed as an ethical matter. The term “asymptomatic” puts the blame on the virus and helps to dispense social failures. In contrast, “undetected” places the burden on society and should help to fix attention in what we can do better. Thus, if we are forced to err because of incomplete information the way we err must be ethically considered.

## Data Availability

The data that support the findings of this study are available from the Public Health Ministry of Argentina. The data is available under the 'public information law' of Argentina (Ley 27275). The program code is available from github: https://github.com/MNatiello/covid-small

## Acknowledgements

HGS acknowledges support from the University of Buenos Aires. MAN acknowledges support from Kungliga Fysiografiska Sällskapet i Lund (2018-2020). We thank the members of the “Red de modelización de enfermedades infecciosas” (CONICET, Argentina) for valuable discussions, and particularly Verónica Simoy for help in tracking public policy in Argentina and Ignacio Simoy who has helped us to analyse the database using the *R* language.

## A Criteria of suspicious case in Argentina

The early evolution of the criteria (we omit the specifications for sanitary operators) for a case to be suspicious in Argentina is as follows (we indicate correspondence with Italy’s resolutions):

29 February 2020 (Corresponds to Italy’s January resolution) There are two ways to be considered a suspected case:

1. The person has fever and signs of respiratory infection (cough, difficulty in breathing) and a requirement for hospitalisation and no other aetiology that fully explains the clinical presentation and a history of travel or residence in mainland China in the 14 days prior to the onset of symptoms.
2. The person has fever and signs of respiratory infection (cough, odynophagia, difficulty in breathing) and either a history of travel or residence in the province of Hubei (China) in the 14 days prior to the onset of symptoms or known close contact with a probable or confirmed case of COVID-19 infection, in the 14 days prior to the onset of symptoms or exposure in a health centre in a country where confirmed cases of COVID-19 have been attended, in the 14 days prior to the onset of symptoms or visited or worked in a live animal market in any city in China.

–

5 March 2020(Corresponds to Italy’s late February resolutions) The person has fever and one or more respiratory symptoms (cough, difficulty in breathing, odynophagia) without another aetiology that fully explains the clinical presentation, and that in the last 14 days either has been in contact with confirmed or probable COVID-19 cases or has a history of travel or presence in areas with local transmission of SARS CoV-2

–

16 March 2020 (Corresponds approximately to Italy’s 9 March resolution) In addition to the previous situation, a new suspected case begins to be considered: Any person with severe acute respiratory disease who requires mechanical ventilation due to their respiratory symptoms, without other aetiology that explains it, even without epidemiological link.

–

21 March 2020

Travel history to specific countries is substituted by travel abroad.

Specifications are given for severe acute respiratory disease, defined as pneumonia and one of the following:

Respiratory rate:> 30 / min,

Sat O2 <93% (ambient air),

Mechanical assistance requirement,

Increase in infiltrates> 50% in 24-48 hours,

Altered consciousness,

CURB-65 ≥2 points,

Unit of Intensive Therapy requirement,

and without another aetiology that explains the clinical picture.

–

30 March 2020

The issue of travel or residence in areas of local transmission (either community or by conglomerates) of COVID-19 in Argentina is added. In the case of pneumonia, the other conditions raised on 21 March are no longer required.

–

8 April 2020

Cases added:

Health workers presenting fever and and one symptom among (cough, odynophagia or difficulty in breathing)

–

16 April 2020

To the requirements raised on 5 March, in the case of symptoms that could accompany fever, the following is added: anosmia/dysgeusia.

–

13 May 2020

Anyone presenting fever (37.5ºC or more) and one or more of (cough, odynophagia, difficulty in breathing, anosmia, dysgeusia) of recent presentation, without another clinical explanation AND precedents of travelling to (or residence in) places with viral circulation or contact with confirmed cases of COVID-19 Anyone presenting anosmia/dysgeusia is to be observed by 72hs and then tested. Health workers with two or more of the described symptoms.

6 June 2020

Anyone presenting two or more of (fever −37.5°C or more-, cough, odynophagia, difficulty in breathing, anosmia, dysgeusia) AND (having being present in a zone with viral circulation OR residing in a “popular neighbourhood”-slum-) OR requiring hospitalization) Health workers and any with close contact with a COVID-19 case presenting ate least one symptom.

–

4 August 2020

Three symptoms added to the set: headache and vomits and diarrhoea.

–

11 September 2020

Added symptom: myalgia

Anyone presenting fever (37.5°C or more) and one or more of (cough, odynophagia, difficulty in breathing, anosmia, dysgeusia) of recent presentation, without another clinical explanation. Anyone presenting anosmia/dysgeusia. For health workers and inhabitants of “popular neighbourhoods” the requisite is of one symptom.

## Footnotes

1 For an epistemological discussion in the context of COVID-19 the reader may consult [(work in press) 10].

2 Extrinsic stochasticity misses the root of the stochastic phenomena[8, 9, 20]

3 [Pub Med’s website](https://pubmed.ncbi.nlm.nih.gov/) accessed 2020-08-28

4 [resolution 27 January](https://www.trovanorme.salute.gov.it/norme/renderNormsanPdf%3Fanno%3D2020%26codLeg%3D72847%26parte%3D1%2B%26serie%3Dnull), accessed 2020-08-26

5 [resolution 22 February](http://www.siditalia.it/pdf/coronavirus/doc_ministero%20salute.pdf) accessed 2020-08-26

6 [resolution 9 March](https://www.trovanorme.salute.gov.it/norme/renderNormsanPdf?anno=2020&codLeg=73669&parte=1%20&serie=null) accessed 2020-08-26

7 [resolution 31 January](https://www.ordinemedicipa.it/pdf/1600_-1.-1) accessed 2020-08-26

8 [resolution 27 February](https://www.trovanorme.salute.gov.it/norme/renderNormsanPdf?anno=2020&codLeg=73448&parte=1%20&serie=null) accessed 2020-08-26

9 [resolution 29 May](https://www.trovanorme.salute.gov.it/norme/renderNormsanPdf%3Fanno%3D2020%26codLeg%3D74178%26parte%3D1%20%26serie%3Dnull) accessed 2020-08-26

10 The Argentine Ministry of Health provides on a daily basis an anonymized copy of the data set corresponding to the nation-wide reported cases in epidemic outbreak for the National Science Council (CONICET).

11 There are 493 cases with reported symptoms where none of the symptoms match the HS expectation.

12 Report of symptoms is not an obligation for the sanitary units.

13 Recommendation of 13 March 2020 (in Swedish) [General information (English)](https://www.folkhalsomyndigheten.se/the-public-health-agency-of-sweden/communicable-disease-control/covid-192/protect-yourself-and-others-from-spread-of-infection/) accessed 2020-10–13.

14 Data available at buenosaires.gob.ar/datosabiertos

15 The accuracy of serological tests used to evaluate *a-posteriori* the prevalence of COVID-19 can be questioned based on works that indicate that not all people presenting immunological reactions to the virus develop humoral antibodies [30, 31, 22]

## Notes

### Competing Interest Statement

The authors have declared no competing interest.

### Funding Statement

Kungliga Fysiografiska Sallskapet (M. Natiello)

